# Disruptions of Hierarchical Cortical Organisation in Early Psychosis and Schizophrenia

**DOI:** 10.1101/2023.05.02.23289376

**Authors:** Alexander Holmes, Priscila T. Levi, Yu-Chi Chen, Sidhant Chopra, Kevin M. Aquino, James C. Pang, Alex Fornito

**Affiliations:** Turner Institute for Brain and Mental Health, School of Psychological Sciences, Monash University, Melbourne, Australia; Brain and Mind Centre, University of Sydney, Sydney, Australia; Department of Psychology, Yale University, New Haven, USA; School of Physics, University of Sydney, Sydney, Australia

## Abstract

**Background:** The cerebral cortex is organised hierarchically along an axis that spans unimodal sensorimotor to transmodal association areas. This hierarchy is often characterised using low-dimensional embeddings, termed gradients, of inter-regional functional coupling estimates measured with resting-state functional magnetic resonance imaging (fMRI). Such analyses may offer insights into the pathophysiology of schizophrenia, which is frequently linked to dysfunctional interactions between association and sensorimotor areas.

**Methods:** To examine disruptions of hierarchical cortical function across distinct stages of psychosis, we applied diffusion map embedding to two independent fMRI datasets: one comprised 114 patients with early psychosis and 48 controls, and the other comprising 50 patients with established schizophrenia and 121 controls. We then analysed the primary sensory-fugal and secondary visual-to-sensorimotor gradients of each participant in both datasets.

**Results:** There were no significant differences in regional gradient scores between patients with early psychosis and controls. Patients with established schizophrenia showed significant differences in the secondary, but not primary, gradient relative to controls. Gradient differences in schizophrenia were characterised by lower within-network dispersion in the Dorsal Attention (p_FDR_<.001), Visual (pFDR=.003), Frontoparietal (pFDR=.018), and Limbic (pFDR=.020) networks and lower between-network dispersion between the Visual network and other networks (pFDR<.001).

**Conclusions:** These findings indicate that differences in cortical hierarchical function occur along the secondary visual-to-sensorimotor axis rather than the primary sensory-fugal axis, as previously thought. The absence of differences in early psychosis suggests that visual-sensorimotor abnormalities may emerge as the illness progresses.

## Introduction

Converging evidence from tract-tracing, lesion, and physiological studies indicates that cortical systems are organised along a hierarchical axis, anchored by unimodal sensorimotor areas at one end and transmodal association areas at the other (1,2). Information-processing along this hierarchy is thought to allow the integration of incoming sensory signals with a diverse range of multimodal interoceptive and exteroceptive signals (3,4). Accordingly, disturbances of this hierarchy are thought to negatively impact cognitive and clinical outcomes (5–9).

The symptoms of psychosis are thought to arise from impaired brain connectivity and a dissolution of integrative, higher-order cognitive processes (10–12). Both sensory deficits and impairments in executive control and attention have been identified in patients, suggesting that a disruption of hierarchical signalling between cortical systems may contribute to disease pathophysiology (13–18).

The hierarchical organisation of cortical function can be investigated using various dimensionality reduction techniques, which decompose estimates of inter-regional functional coupling (FC) measured with resting-state functional magnetic resonance imaging (fMRI) into orthogonal axes of variation (1,7). These axes define smoothly varying *gradients* of regional FC profiles, ordered by the amount of variance in FC similarity that they explain (19–21). The dominant (primary) gradient explains the most variance and represents an axis spanning from visual and other sensorimotor areas to the default mode network (DMN) and other transmodal regions (1,22), consistent with classical descriptions of the sensory-fugal axis of cortical organisation (3,23). The secondary gradient represents an axis spanning from sensorimotor areas to visual areas, differentiating unimodal areas that are situated at approximately the same level in the cortical hierarchy (3).

Recent work suggests that patients with established schizophrenia show a contraction of the primary sensory-fugal gradient (24), suggesting reduced differentiation between transmodal and sensorimotor areas. However, whether the secondary, sensorimotor-visual gradient is also affected by the illness remains unknown. Moreover, since this study investigated patients with established schizophrenia, it is unclear whether changes in hierarchical cortical organisation are apparent from the earliest stages of the disease or emerge only with prolonged illness. To address these questions, we investigated the organisation of the cortical hierarchy by comparing the primary and secondary FC gradients of participants in two independent cohorts––one with patients in the early stages of psychosis and another with patients with established schizophrenia––allowing us to determine whether disrupted hierarchical function persists across different illness stages. Given the prominent role ascribed to disruptions of hierarchical processing in the onset of psychotic symptoms (13,14,18), we hypothesised that both cohorts of early psychosis and schizophrenia will show disrupted organisation of the dominant sensory-fugal gradient, characterised by a reduced functional differentiation between sensorimotor and transmodal association areas.

## Methods

### Participants

We used open-access neuroimaging and behavioural data from 178 participants from the Human Connectome Project for Early Psychosis (HCP-EP) dataset (57 controls; 121 early psychosis patients) (25) and 171 participants from the UCLA Consortium of Neuropsychiatric Phenomics (CNP) dataset (121 controls; 50 schizophrenia patients) (26). See Tables 1-3 for demographic and clinical information across both datasets.

**Table 1:** Demographic information for controls and patients within the HCP-EP and CNP datasets.

|  | HCP-EP |  | CNP |  |
| --- | --- | --- | --- | --- |
|  | Controls | Early Psychosis Patients | Controls | Schizophrenia Patients |
| Mean age, years (*SD*) | 24.79 (3.95) | 22.77 (3.75) | 31.47 (8.75) | 36.46 (8.88) |
| Sex male, n (%) | 40 (68.97%) | 79 (65.29%) | 65 (53.72%) | 38 (76.00%) |
| Mean illness duration, years (*SD*) |  | 1.83 (1.35) |  | Not available |
| Medication naïve, n (%) |  | 67 (55.37%) |  | 6 (12.00%) |
| Chlorpromazine Equivalence, mg/d (*SD*) |  | 157.96 (228.11) |  | 224.50 (449.25) |

**Table 2:** Mean scores across clinical measures of patients within the HCP-EP cohort.

| Clinical Measure | Subscale | Mean (*SD*) |
| --- | --- | --- |
| Positive and Negative Syndrome Scale | Positive Symptoms | 14.10 (4.75) |
|  | Negative Symptoms | 12.56 (5.58) |
|  | Cognitive Symptoms | 10.84 (3.01) |

**Table 3:**
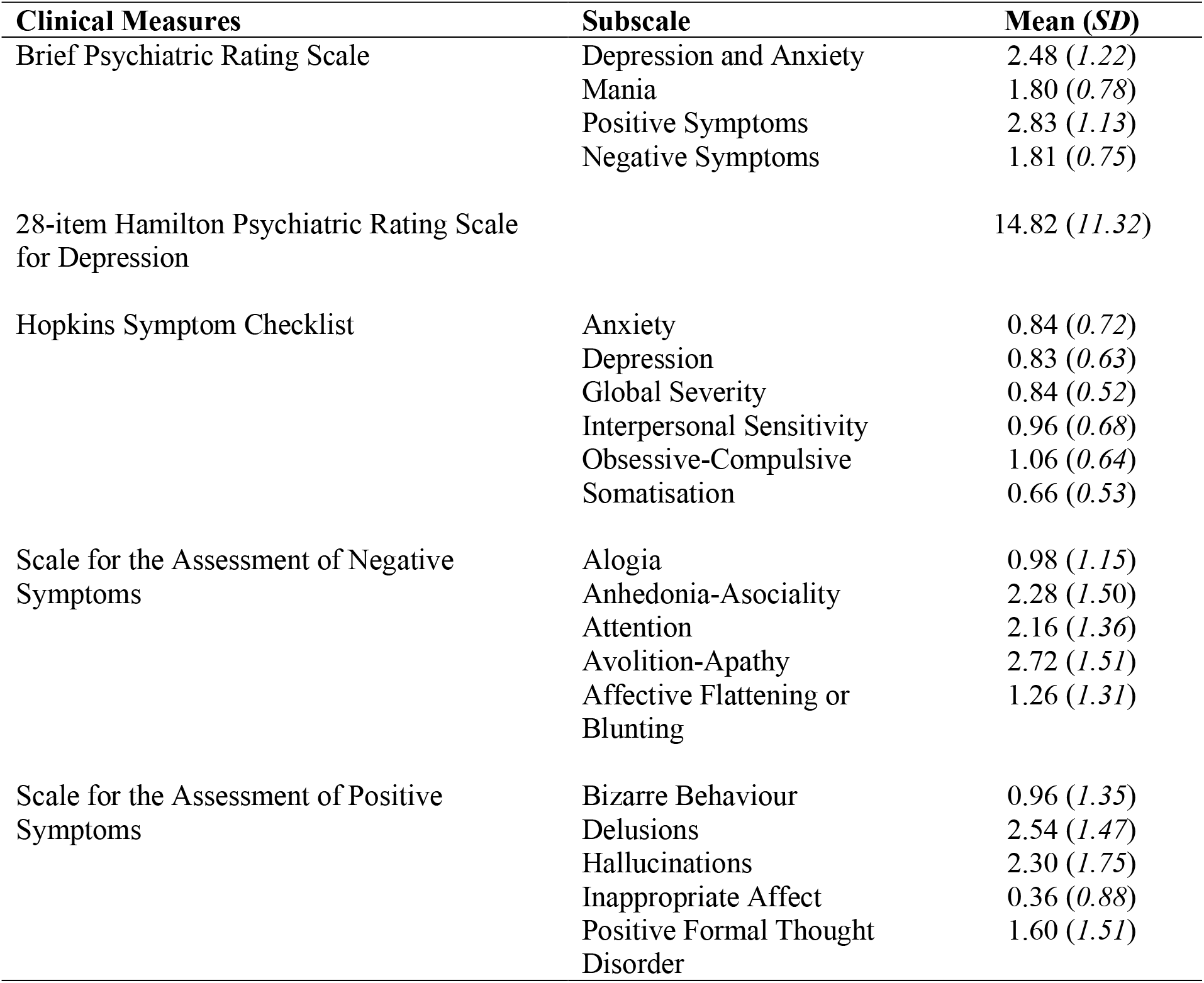
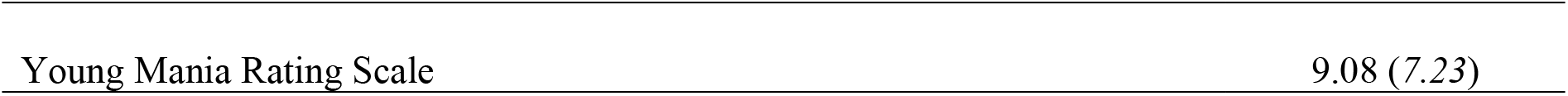
Mean scores across clinical measures of patients within the CNP schizophrenia cohort.

Early psychosis patients in the HCP-EP sample were assessed with the Structured Clinical Interview for DSM-5: Research Version (SCID-5-RV) (27) to confirm diagnoses of either non-affective (schizophrenia, schizophreniform, schizoaffective, psychosis NOS, delusional disorder, brief psychotic disorder) or affective psychosis (major depression with psychosis or bipolar disorder with psychosis). Patients must have experienced onset of psychosis within five years of recruitment. Healthy controls did not meet criteria for a diagnosis of any anxiety or psychotic disorder, were not taking psychotropic medication, had no first-degree relative with schizophrenia spectrum disorder, and had no history of psychiatric hospitalisation. Schizophrenia patients in the CNP sample were diagnosed with the Structured Clinical Interview for DSM-IV (SCID-IV) (28). Healthy controls did not have a history of psychotic or substance use disorder, and no current diagnosis of mood or anxiety disorders. Further recruitment details, inclusion criteria, exclusion criteria, and participant screening details can be found in the original HCP-EP (25) and CNP papers (26).

### MRI Data Acquisition and Image Preprocessing

#### Human Connectome Project for Early Psychosis (HCP-EP) dataset

Imaging data for the HCP-EP dataset were acquired on 3T Siemens MAGNETOM Prisma scanners at the Brigham and Women’s Hospital, McLean Hospital, and Indiana University School of Medicine (25). Acquisition parameters between sites were harmonised using Siemens-specific QA tools, phantom measurements (fBIRN and NIST phantoms), and traveling human subjects. Anatomical T1-weighted (T1w) MPRAGE images (0.8 mm isotropic voxels, 2400 ms TR, 2.22 ms TE) and two sessions of multiband resting-state echo-planar images (EPI) were acquired with AP and PA phase encoding directions (420 volumes, 2 mm isotropic voxels, multiband factor=8,800 ms TR, 37 ms TE), resulting in a total of four resting-state runs per individual.

Raw data were downloaded from the NIMH Data Archive and pre-processed on the MASSIVE high-performance computing facility (29) in accordance with HCP pre-processing pipelines (30). Basic pre-processing was conducted using FMRIB Software Library (FSL), which included skull striping of T1w and fMRI via FSL BET, image co-registration via FSL FLIRT, and removal of the first 10 functional volumes to account for initial inhomogeneity in fMRI signals. Fieldmap correction using EPI scans with opposite phase encoding directions was applied using FSL’s topup function and fine-tuned using FreeSurfer’s bbregister function to account for spatial distortion and static field inhomogeneities. Imaging data were nonlinearly registered to the MNI152 space with 2 mm resolution via FSL FNIRT. Functional data were denoised via a temporal high-pass filter of 2000 alongside FSL MELODIC, which uses Independent Component Analysis (ICA) to decompose the data into distinct components corresponding to either signal or noise, whereby FSL-FIX (31,32) removed structured noise components from the functional timeseries using pre-trained weights from the HCP_hp2000 classifier.

Motion during fMRI acquisition was quantified by measuring framewise displacement (FD) according to the Jenkinson method (33). 11 participants were excluded based on the following criteria: mean FD > 0.25 mm across all timeseries volumes, FD > 0.2 mm in more than 20% of the volumes, or FD > 5 mm in any volume. Additionally, 5 participants were excluded due to incomplete imaging or diagnostic data. The final HCP-EP sample included 162 participants (48 controls; 114 early psychosis patients).

#### UCLA Consortium of Neuropsychiatric Phenomics (CNP) dataset

Imaging data for the CNP dataset were acquired on 3T Siemens Trio scanners at the Ahmanson-Lovelace Brain Mapping Center and Staglin Center for Cognitive Neuroscience at the University of California, Los Angeles (26). Anatomical T1w MPRAGE images (1 mm isotropic voxels, 1900 ms TR, 2.26 ms TE) and resting-state EPI (152 volumes, 4 mm isotropic voxels, 2000 ms TR, 30 ms TE) were acquired for each individual. Image pre-processing of the CNP dataset was conducted as described elsewhere (34). In short, the fMRIprep v1.1.1 pipeline (35) was used to pre-process the fMRI data for each participant, which included spatial normalisation, slice-timing correction, and realignment to a reference volume via MCFLIRT. Denoising was performed using ICA-AROMA(36), which shows good performance in mitigating motion-related artifact (37).

### Functional Connectivity (FC) Matrices

The pre-processed individual functional data from both datasets were projected onto the individual’s cortical surface and parcellated into 1000 discrete regions across both hemispheres using the Schaefer atlas (38). The atlas is most appropriate for FC analyses and assigns each region to the 7 Yeo large-scale functional networks; Visual, Somatomotor, Dorsal Attention (DAN), Ventral Attention (VAN), Limbic, Frontoparietal, and Default Mode networks (DMN) (2). Region-to-region FC matrices were constructed by calculating pairwise Pearson product-moment correlations of regional timeseries data.

### Functional Gradient Analyses

FC matrices from both datasets were z-transformed and thresholded to only include the top 10% of connections at each row of the matrix, as done previously (1). Diffusion map embedding (19,20) was implemented through the Python-based BrainSpace toolbox (21), which identifies a low-dimensional manifold underlying the FC matrices. Briefly, the method first involved obtaining the cosine similarity in FC profiles between pairs of parcels. Eigendecomposition on a diffusion probability matrix estimated from the similarity matrix was then used to obtain functional gradients of the FC data (eigenvectors) and the variance explained by each gradient (related to the eigenvalues). Figure 1 further describes the pipeline to obtain FC and gradient data for each individual.

**Figure 1:**
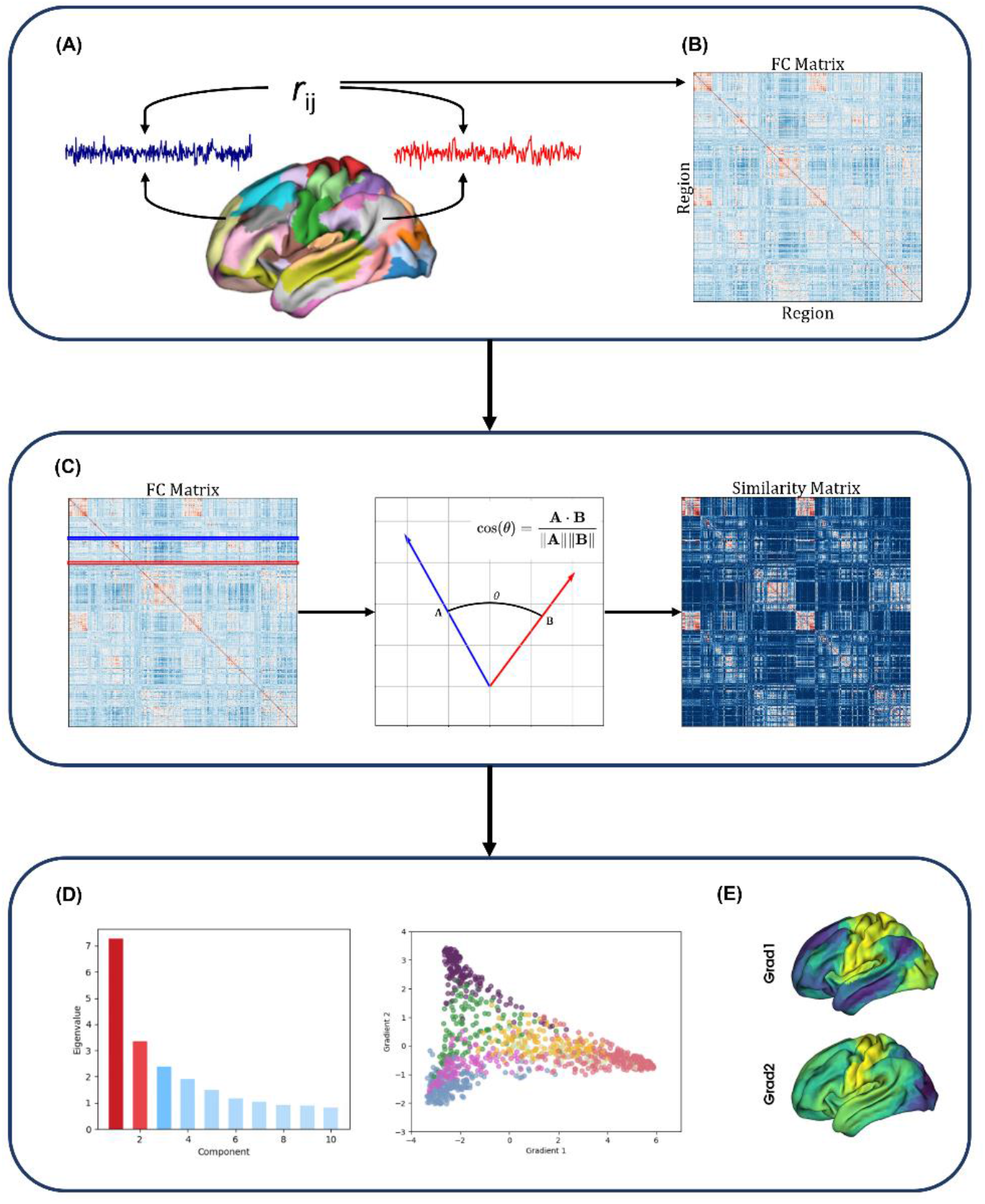
The functional connectivity and functional gradient pipeline. (A) Functional timeseries data are extracted from preprocessed fMRI scans and parcellated into 1000 regions-of-interest using the Schaefer atlas. (B) Pearson product-moment correlations are calculated between each pair of regional timeseries to produce a 1000*1000 FC matrix. (C) Similarity in connectivity profiles between regions within the FC matrix are estimated based on the difference in angle between pairs of vectors of the FC matrix in a 1000-dimensional space. Similarity between each possible pair of regions is then expressed through a cosine similarity matrix. (D) The application of diffusion map embedding returns a series of eigenvectors ordered by decreasing eigenvalues. The first two eigenvectors are kept and correspond to the primary and secondary gradient. (E) These eigenvectors are plotted on the cortical surface to visualise the spatial topology of each gradient.

The spatial organisation of gradients can vary across individuals. Moreover, since gradient scores are inherently unitless, they are not directly comparable between individuals. We therefore used joint embedding to align each participant’s gradients with respect to a common template to allow comparison across individuals (39). A separate template was generated for each dataset using a group-average FC matrix obtained from an independent cohort of 40 controls. Briefly, the joint embedding procedure concatenated an individual’s similarity matrix, the template similarity matrix, and off-diagonal couplings between the individual and template matrices. This joint similarity matrix was then used as the input for diffusion map embedding. For computational efficiency, we performed the procedure separately for each participant, while still ensuring that the aligned individual-specific gradients existed within a common coordinate space.

We only analysed the first two dominant gradients (primary and secondary gradients) as they have been most extensively studied in the literature and map onto clearly defined axes of cortical organisation (1,7). To determine whether whole-brain gradient architecture differed between controls and patients in the HCP-EP and CNP datasets, we used general linear models to examine differences in regional gradient scores between patients and control, using, age, sex, and scan site as covariates. Post-hoc effect sizes were obtained via partial correlations after controlling for these covariates and Benjamini-Hochberg False Discovery Rate (FDR) corrections were applied to control for multiple comparisons.

We also analysed gradient scores at the 7 Yeo functional network level (2) using within-network dispersion and between-network dispersion metrics (40). These metrics were calculated for each participant based on the Euclidean distance between points in a two-dimensional space with coordinate axes representing the aligned primary and secondary gradient scores. Within-network dispersion calculates the Euclidean distance between all regions within a single network and the network’s centroid, with lower values suggesting more uniform patterns of FC within the network. Between-network dispersion calculates the Euclidean distance between pairs of network centroids across different networks, with lower values suggesting less differentiation of the FC patterns associated with nodes in different networks.

### Behavioural Correlates and Clinical Outcomes

We used canonical correlation analysis (CCA) in the CNP dataset to examine the association between gradient dispersion metrics and clinical symptoms, assessed using 22 different behavioural scales (Table 3) (26). To reduce the dimensionality of the variables, we applied principal component analysis (PCA) on the gradient dispersion metrics among patients with schizophrenia, reducing the data from 21 between-network dispersion factors to 6 components that explained 92.54% of the variance in the data. Similarly, PCA was applied on the 22 clinical items to minimise the collinearity between these variables. We retained 10 components that explained 91.33% of the variance in the data.

CCA was applied on the 6 gradient dispersion and 10 clinical measure components, with age and sex included as confounding variables, to identify linear combinations of the data that maximally covary (41). We used a recently developed permutation-based procedure with 50,000 iterations to measure associations between gradient dispersion metrics and clinical outcomes on the resulting canonical variates (42). The p-values of canonical modes were controlled using family-wise error rate (FWER), which is more appropriate than FDR in the context of CCA (42). Robust estimates of canonical loadings were obtained via bootstrapping with 1000 iterations of the correlation between each gradient dispersion and clinical measure, and corresponding canonical variate.

## Results

### Gradient Architecture in Early Psychosis

In the HCP-EP dataset, we observed a primary cortical gradient consistent with the classical sensory-fugal hierarchy and a secondary gradient spanning from visual to other sensorimotor areas for both healthy controls (HC) and early psychosis patients (EP) (Fig. 2A). The variance in FC explained by each gradient did not significantly differ between patients and controls; the first two gradients explained, on average, 38.92% (SD=6.35) and 21.50% (SD=3.04) among controls and 41.12% (SD=5.29) and 21.17% (SD=3.98) among early psychosis subjects, respectively. Compared to controls, early psychosis patients demonstrated no significant differences in regional gradient scores along both the primary and secondary gradients following correction for multiple comparisons (Fig. 2B). Similarly, we observed no significant differences in between- and within-network dispersion metrics between controls and early psychosis patients (Fig. 2C). Medication use (Chlorpromazine Equivalence, mg/d) was not associated with regional gradient scores among patients with early psychosis.

**Figure 2.**
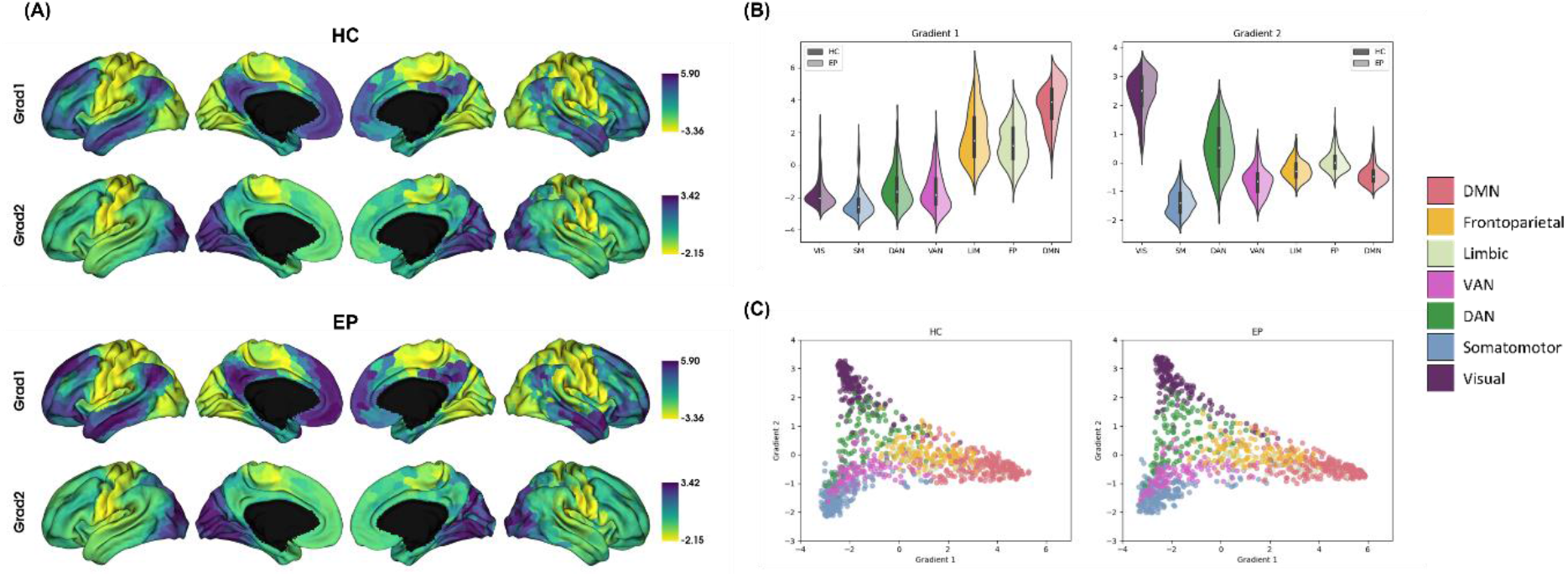
Gradient Architecture in Early Psychosis. (A) Spatial topography of the primary sensory-fugal and secondary visual-to-sensorimotor gradients in healthy controls (top) and early psychosis patients (bottom). (B) Average primary (left) and secondary (right) gradient scores along the 7 Yeo networks. Healthy controls are displayed on the left side of each violin plot (darker shade), while early psychosis patients are displayed on the right (lighter shade). (C) Average gradient scores of healthy controls (HC; left) and early psychosis patients (EP; right) on a two-dimensional coordinate gradient space. The colours represent the 7 Yeo networks. Each point represents a cortical region.

### Gradient Architecture in Established Schizophrenia

In the CNP dataset, we also observed primary and secondary gradients that follow the sensory-fugal and visual-to-sensorimotor axes, respectively (Fig. 3A). As in the HCP-EP dataset, the variance explained by each gradient did not significantly differ between patients and controls, with the first two gradients explaining 30.52% (SD=3.99) and 21.33% (SD=2.73) among healthy controls and 29.27% (SD=4.86) and 20.30% (SD=3.06) among schizophrenia patients, respectively. Compared to controls, schizophrenia patients demonstrated no significant differences in regional scores along the primary gradient but showed significantly lower scores in the visual network along the secondary gradient (pFDR<.05) (Fig. 3B). Statistical differences in the secondary gradient displayed on the cortical surface are shown in Fig. 3D, including effect sizes for significant regions. Medication use (Chlorpromazine Equivalence, mg/d) was not associated with regional gradient scores among patients with schizophrenia.

**Figure 3.**
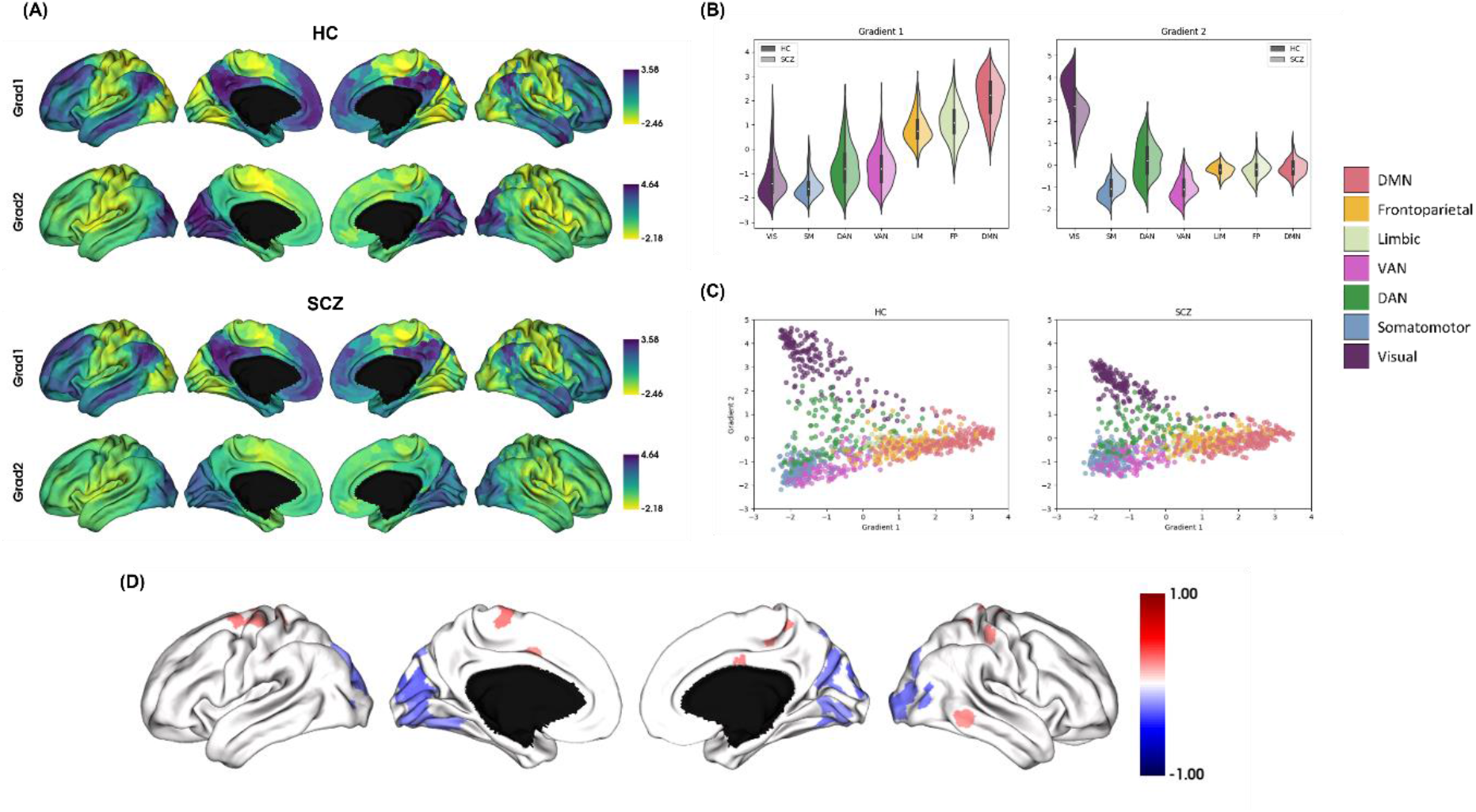
Gradient Architecture in Schizophrenia. (A) Spatial topography of the primary sensory-fugal and secondary visual-to-sensorimotor gradients in healthy controls (top) and schizophrenia patients (bottom). (B) Average primary (left) and secondary (right) gradient scores along the 7 Yeo networks. Healthy controls are displayed on the left side of each violin plot (darker shade), while schizophrenia patients are displayed on the right (lighter shade). (C) Average gradient scores of healthy controls (left) and schizophrenia patients (right) on a two-dimensional coordinate gradient space. The colours represent the 7 Yeo networks. Each dot represents a cortical region. (D) Pearson correlation coefficient effect sizes of regions that significantly differed along the secondary gradient between healthy controls and schizophrenia patients.

Analysis of network-level gradient dispersion along the secondary gradient revealed significantly lower within-network dispersion in the Visual (r_PARTIAL_=-0.225, p_FDR_=.012), Dorsal Attention (r_PARTIAL_=-0.293, p_FDR_ < .001), Limbic (r_PARTIAL_=-0.179, p_FDR_=.035), and Frontoparietal (r_PARTIAL_=-0.182, p_FDR_=.035) networks, alongside significantly lower between-network dispersion between the Visual network and all other networks, including the Somatomotor (r_PARTIAL_=-0.291, p_FDR_=.001), Dorsal Attention (r_PARTIAL_=-0.283, p_FDR_=.001), Ventral Attention (r_PARTIAL_=-0.274, p_FDR_=.001), Limbic (r_PARTIAL_=-0.241, p_FDR_=.006), Frontoparietal (r_PARTIAL_=-0.279, p_FDR_=.001), and DMN (r_PARTIAL_=-0.255, p_FDR_=.004) (Fig 3C).

### Gradient Dispersion and Clinical Outcomes in Established Schizophrenia

The CCA revealed a single statistically significant canonical dimension (CCA r=0.77; p_FWER_=.028; Fig. 4A). We found significant positive loadings of the between-network dispersion pairs of the Visual and DAN, Visual and Frontoparietal, Visual and DMN, Somatomotor and VAN, Somatomotor and Frontoparietal, Somatomotor and DMN, DAN and Frontoparietal, VAN and Frontoparietal, and VAN and DMN (Fig. 4B). The strongest loadings were observed on pairings of higher-order and lower-order networks, such as the Frontoparietal and Somatomotor networks or DMN and Somatomotor network. Such pairings mirror the regional anchors of the primary sensory-fugal gradient. Notably, the between-network pair of the Visual and Somatomotor networks did not significantly load onto the canonical mode, where the distance between these unimodal networks most aligns with the eccentricity of the secondary visual-to-sensorimotor gradient. Clinical variables with significant positive loadings on this dimension included BPRS – Negative Symptoms and SANS – Affective Flattening or Blunting. Clinical variable with significant negative loadings included BPRS – Mania, HSC – Global Severity, HSC – Interpersonal Sensitivity, HSC – Somatisation, SAPS – Delusions, SAPS – Positive Formal Thought Disorder, and YMRS – Total score factors (Fig. 4C).

**Figure 4.**
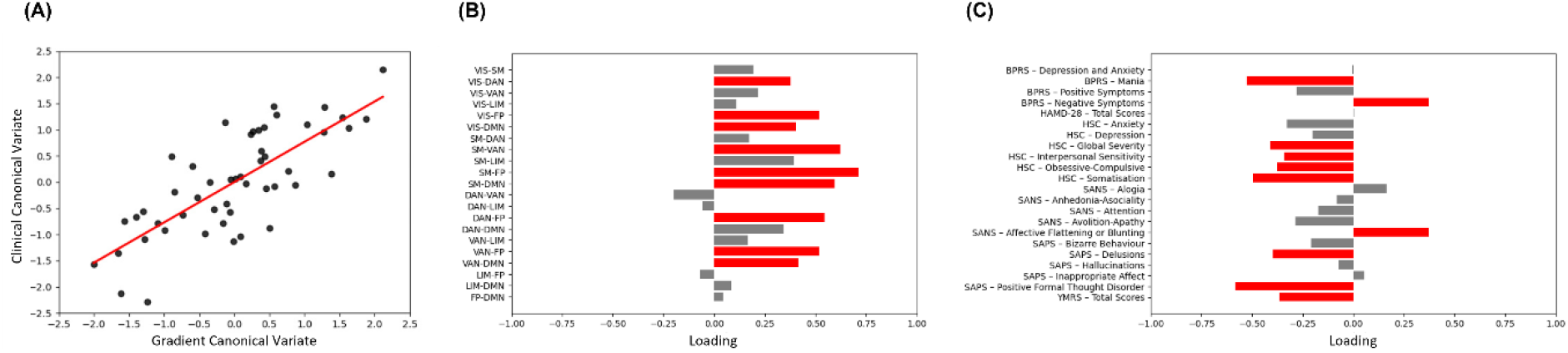
Results of Canonical Correlation Analysis. (A) Association between the gradient canonical variate and clinical canonical variate, with the least-squares regression line plotted in red. (B) Correlations between the original gradient dispersion metrics and the gradient canonical variate. Significant loadings are plotted in red, while non-significant loadings are plotted in grey. (C) Correlations between the original behavioural measures and the behavioural canonical variate. Significant loadings are plotted in red, while non-significant loadings are plotted in grey.

Thus, our findings identify strong associations between the aforementioned between-network gradient dispersion pairs and clinical outcomes in schizophrenia, with increased between-network dispersion associated with elevated scores on clinical measures relating to negative symptoms and flattened affect and decreased between-network dispersion associated with elevated scores on clinical measures relating to positive and general symptoms, including mania, somatisation, delusions, and positive formal thought disorder.

## Discussion

Disturbed connectivity between higher-order and sensorimotor systems is commonly implicated in the genesis of psychotic symptoms (13,14,24). Here, we used diffusion map embedding to characterise gradients of sensory-fugal and visual-to-sensorimotor organisation of the cerebral cortex in patients with early psychosis and established schizophrenia. Contrary to our hypothesis, we found that the primary sensory-fugal gradient was largely unaffected in both early psychosis and schizophrenia patients. The only disruptions we identified affected the secondary visual-to-sensorimotor gradient in schizophrenia, such that visual regions along the gradient showed a reduced differentiation from opposing sensory and motor areas. This reduced differentiation along the secondary visual-to-sensorimotor gradient was not observed in early psychosis. Together, these results suggest that schizophrenia is associated with a blurring of the functional boundaries between different sensory and motor abnormalities that is apparent only after prolonged illness.

### Primary and Secondary Axes of Cortical Organisation in Schizophrenia

Evaluation of the average gradients within each cohort showed broad similarities in the spatial patterning of the primary and secondary gradients, indicating that neither early psychosis nor schizophrenia are associated with a large-scale reorganisation of cortical function. Studies in other disorders have demonstrated similarly consistent primary and secondary gradient architecture between patients and healthy controls (43,44). This result implies that neurodevelopmental disturbances of brain wiring in schizophrenia are likely subtle, and do not lead to major changes in the dominant modes of cortical activity, even where there are significant group differences along the gradient.

The contraction of the visual-to-sensorimotor gradient observed in schizophrenia patients suggests that the disorder preferentially affects this secondary gradient, rather than the primary sensory-fugal axis. Although the primary sensory-fugal gradient is typically associated with interactions between unimodal and higher-order transmodal associative processes, deviations along the secondary visual-to-sensorimotor gradient may potentially be related to schizophrenia symptoms tied to perceptual disturbances, such as hallucinations. Within the context of Mesulam’s hierarchical framework (3), the visual-to-sensorimotor axis differentiates between unimodal sensory regions occupying the same synaptic level upstream along the hierarchy. Our findings suggest that different unimodal systems in schizophrenia are uniquely less differentiated, suggesting a blurring of the boundaries between different sensory modalities.

Changes in the secondary gradient of schizophrenia patients subsequently impacts macroscale network organisation, as evidenced by reductions in between-network dispersion between the Visual and other networks. This contraction in gradient scores indicates that the FC patterns differentiating the Visual network from other unimodal sensorimotor networks are less pronounced in schizophrenia. The spread of gradient scores in schizophrenia patients was also disturbed within individual networks, evidenced by reductions in within-network dispersion in the Visual, DAN, Limbic, and Frontoparietal networks. Notably, the greatest effect was observed in the Visual network and DAN, such that regions in these networks have more homogenous FC profiles.

Our findings contrast with a prior study reporting disruptions of the first principal gradient in schizophrenia patients (24). This discrepancy may arise from differences in clinical stage and severity between samples. While it is difficult to directly compare the severity of symptoms between these two samples due to the different clinical measures used, schizophrenia patients within the present sample scored higher on domains assessing positive symptoms than negative symptoms compared to the previous study (24). As negative symptoms are closely associated with disrupted cognition, such symptoms may predict deviations along the sensory-fugal hierarchy, given the link between cortical hierarchical organisation and the propagation of higher-order thought (4,45).

### Gradient Architecture in Early Psychosis

The absence of any gradient differences in early psychosis suggests that abnormalities along the visual-to-sensorimotor axis may only emerge as the illness progresses. This may be due to heterogeneity within early-phase psychosis and the greater severity of brain changes associated with later stages of schizophrenia (46–48). Clinical outcomes are more varied in early psychosis, with 42% of individuals with early psychosis showing clinical improvement (49). Although previous studies investigating early psychosis found diminished FC (50–54), other research suggests that individuals with early psychosis possess similar patterns of connectivity to healthy controls (55), which aligns with the gradient architecture in early psychosis observed in the present study. One possibility is that local edge-level differences are more sensitive to a patient state, and thus pick up on brain activity patterns related to present symptoms. Gradient-based approaches may instead reflect trait-level global differences which are not readily modified by active symptoms, being more stable and less altered in disease. Thus, while edge-level FC may be affected in early psychosis, the changes may not be severe enough to produce widespread changes in cortical organisation.

### Clinical Correlates of Disrupted Cortical Organisation

More severe schizophrenia symptoms in the CNP cohort were associated with atypical gradient architecture. Specifically, reduced network dispersion between most network pairs was associated with higher scores on clinical measures assessing general and positive symptoms, such as delusions. In contrast, elevated between-network dispersion was associated with higher scores on measures assessing negative symptoms, such as affective flattening and blunting.

The between-network dispersion values between pairs of higher-order and lower-order networks showed the strongest loadings of gradient metrics within the CCA (e.g., between the Visual network and DMN or Somatomotor network and DMN). Interestingly, these pairings mirror the regional anchors of the primary sensory-fugal gradient, rather than the secondary visual-to-sensorimotor gradient. Although the present group-level analyses suggest that the greatest shift in gradient architecture between healthy controls and individuals with schizophrenia occurs along the secondary gradient, dispersion metrics assess gradient architecture within a multidimensional space, incorporating information from both the primary and secondary gradients (40). Past studies investigating brain-behaviour relationships in schizophrenia have consistently linked symptoms with the functions of transmodal association areas (56–58); however, the complexity of these cognitive and behavioural processes often necessitates coordinated function from multiple hierarchical pathways (13,16,17). Although the present study highlights the relative significance of visual-to-sensorimotor functioning between individuals with schizophrenia and healthy controls, symptom profiles may still be mediated by deviations along multiple axes of organisation (59).

### Limitations

This study used two separate datasets to investigate gradient differences in early psychosis and schizophrenia; hence, no direct comparisons could be made between the two patient groups. We could only examine how each patient group differed with respect to their own control group. Thus, while these findings are suggestive of a progression of gradient disruptions with illness, caution is warranted when drawing longitudinal conclusions from cross-sectional data. Differences between the two cohorts could be attributable to differences in clinical severity, medication exposure, and numerous other clinical and demographic variables. Post-hoc analyses revealed that medication use was not associated with disturbed gradient scores along either the primary or secondary gradient in the schizophrenia group.

Furthermore, the present study investigated cortical gradients at the level of parcellated functional connectivity maps, rather than vertex-wise connectivity. This approach could influence the decomposition of the functional connectivity matrix used in diffusion map embedding, as the geometry of data within the higher-dimensional space may differ between a lower resolution connectivity matrix and a higher resolution connectivity matrix. Such differences could influence the resulting decomposition, producing a disparate set of gradients. However, the spatial distribution of regional gradient scores within the present study was consistent with prior literature (1). Moreover, this decision is unlikely to influence the observed group differences, as the same parcellation was applied to all subjects across groups. Nonetheless, further investigation of such methodological choices, including the use of individually-tailored parcellations (60,61), will be an important avenue for future research.

## Conclusions

We identified novel differences along the secondary gradient of functional connectivity, spanning the visual-to-sensorimotor axis, in individuals with schizophrenia but not early phase psychosis. This result suggests that disturbed cortical organisation in schizophrenia may be related to a reduced differentiation of sensorimotor systems rather than disruptions of hierarchical processing streams. This view is supported by the associations observed between altered organisation of the secondary gradient and a diverse range of clinical rating scales. Our findings thus demonstrate the clinical significance of successive gradients beyond the dominant primary gradient in schizophrenia, suggesting an important role for dysfunction of unimodal systems.

## Data Availability

All data produced are available online at https://www.humanconnectome.org/study/human-connectome-project-for-early-psychosis and https://exhibits.stanford.edu/data/catalog/mg599hw5271

https://exhibits.stanford.edu/data/catalog/mg599hw5271

